# Immunometabolic effects of lactate on B cell function in healthy individuals of different ages

**DOI:** 10.1101/2023.08.07.23293760

**Authors:** Maria Romero, Kate Miller, Andrew Gelsomini, Denisse Garcia, Kevin Li, Dhananjay Suresh, Daniela Frasca

## Abstract

Aging is characterized by chronic systemic inflammation and metabolic changes. When we compared B cells from young and elderly donors, we found that aging induces higher oxygen consumption rates, and especially higher extracellular acidification rates, measures of oxidative phosphorylation and of anaerobic glycolysis, respectively. Importantly, this higher metabolic status, which reflects the age-associated expansion of pro-inflammatory B cell subsets, was found associated with higher secretion of lactate and autoimmune antibodies after in vitro stimulation. B cells from elderly individuals, induce in vitro generation of pro-inflammatory CD4+ T cells from young individuals through metabolic pathways mediated by lactate secretion. Lactate also induces immunosenescent B cells that are glycolytic and express transcripts for multiple pro-inflammatory molecules. These results altogether may have relevant clinical implications and suggest novel targets for therapeutic interventions in patients with inflammatory conditions and diseases.

## Introduction

Lactate, the end product of anaerobic glycolysis, has been considered for long time a discarded metabolic compound, secreted under hypoxic conditions, with various harmful effects. However, increasing evidence indicates that lactate is an active metabolite in cell signalling and has a potent immunoregulatory role, especially at sites of inflammation where its accumulation is a common feature of inflammatory-based diseases and cancer. Lactate effects are due to signaling through its specific receptor G protein-coupled receptor 81, or to transport into cells through monocarboxylate transporters.

In dendritic cells (DCs), lactate has been shown to suppress antigen delivery and presentation as well as antigen degradation ^1^. In the presence of IL-4 and GM-CSF, it inhibits the differentiation of monocytes into DCs ^2^. In tissue-resident macrophages, it enhances TLR4 signaling through MD-2, a co-receptor for TLR4 signaling, leading to NF-κB transcriptional activity, and expression of inflammatory genes ^3^. In CD8+ T cells, it decreases cytotoxic activity, but not cell proliferation, and impairs the secretion of mediators of cytotoxicity ^4^. In CD4+ T cells, lactate induces IL-17 production via nuclear PKM2/STAT3 and synthesis of fatty acids, and leads to the retention of CD4+ T cells in inflamed tissues ^5^ by inhibiting their motility ^6^. In mast cells, lactate inhibits both early and late phases of cell activation in asthma by targeting the receptor MRGPRX2 (MAS-associated G protein-coupled receptor X2) ^7^.

In the context of cancer, lactate promotes tumor growth and metastases ^8, 9^; inhibits differentiation and function of monocytes ^10^; promotes the polarization of tumor-associated macrophages (TAMs) which in turn promotes tumor progression ^11^; induces IL-23 secretion by TAMs, leading to higher local inflammation and increased incidence of tumors ^12^; activates the TH17 but not the TH1 pathway in T cells, also leading to higher inflammation in the tumor microenvironment ^12^; promotes the development and accumulation of MDSCs ^13, 14^ while suppressing the effector function of NK cells, CD4+ T cells, CD8+ T cells and mast cells ^13–15^; decreases the response to checkpoint inhibitors ^16^, as enhances PD-1 expression in highly glycolytic tumors ^17^.

In this study, we have identified for the first time a key role for lactate in the regulation of B cell function and antibody production. We show that lactate induces inflammatory B cells able to express multiple transcripts for markers of the senescence-associated secretory phenotype (SASP) and to secrete autoimmune pathogenic antibodies. Moreover, we show that pharmacologic targeting of lactate enzymes and transporters in B cells might provide a novel approach to target B cell-mediated inflammation. The effects of lactate described herein indeed recapitulate key features of B cells that infiltrate inflammatory tissues like the synovial tissue of Rheumatoid Arthritis (RA) patients, where lactate-induced signaling promotes many pathogenic features and persistence of the B cell infiltrates.

## Methods

### Subjects

Participants were young (30-45 years) and elderly (≥65 years), all recruited at the University of Miami Miller School of Medicine. Participants were healthy and were not taking medications affecting the immune system. Subjects with type-2 diabetes mellitus, autoimmune diseases, congestive heart failure, cardiovascular disease, chronic renal failure, malignancies, renal or hepatic diseases, infectious disease, trauma or surgery, pregnancy, or under substance and/or alcohol abuse were excluded.

All participants signed an informed consent. The study was reviewed and approved by our Institutional Review Board (IRB, protocols #20070481 and #20160542), which reviews all human research conducted under the auspices of the University of Miami. Characteristics of the recruited participants (age, gender, serum metabolic measures) are shown in Supplementary Table 1.

### PBMC collection

Blood was drawn in Vacutainer CPT tubes (BD 362761), then PBMC were isolated and cryopreserved. PBMC were thawed and cultured at the concentration of 1×10^6^/mL in complete medium (c-RPMI) [RPMI 1640 (Gibco ThermoFisher 11875-093), supplemented with 10% FBS (Gibco 10437-028), 100 U/mL Penicillin-Streptomycin (Gibco 15140-122), and 2 mM L-glutamine Gibco 25030-081)]. FBS was certified to be endotoxin-free. After thawing, viability of the PBMC was checked by trypan blue counting and samples were discarded if viability was <75%.

### B cell isolation and in vitro stimulation

B cells were isolated from thawed PBMC by magnetic sorting using CD19 Microbeads (Miltenyi Biotec 130-121-301) according to the MiniMACS protocol (20 µL Microbeads + 80 µL PBS, for 10^7^ cells). Cell preparations were typically >95% pure.

B cells at the concentration of 10^6^/mL of c-RPMI were left unstimulated or were stimulated 2-7 days with 5 µg/10^6^ cells of CpG (ODN 2006 InVivogen). In glycolytic experiments, B cells were stimulated overnight with CpG (5 µg/10^6^ cells), in the absence or presence of lactate (Na-Lactate, SIGMA 98867-56-1, 10 mM/10^6^ cells).

To evaluate RNA expression of molecules involved in metabolic pathways, the mRNA was extracted from unstimulated or stimulated B cells with µMACS mRNA isolation kit (Miltenyi Biotec), eluted into 75 µL of pre-heated (65°C) elution buffer, and stored at -80°C until use.

To evaluate the expression of inflammatory markers, unstimulated or stimulated B cells were resuspended in TRIzol (Ambion) (10^6^ cells/100 µl), then RNA extracted for qPCR. Total RNA was isolated according to the manufacturer’s protocol, eluted into 10 µl of pre-heated elution buffer and stored at -80°C until use.

Culture supernatants were collected at day 2 of CpG stimulation to measure the secretion of pro-and anti-inflammmatory cytokines by BD^TM^ Cytometric Bead Array, CBA (BD 560484) human TH1/TH2/TH17 kit, or lactate using the L-lactate assay kit (Cayman Chemicals 700510), following manufacturer’s instructions. Day-7 supernatants were evaluated for autoimmune IgG antibodies by ELISA (MyBioSource MBS390120).

### B cell:CD4+ T cell co-cultures

B cells, sorted from young or elderly individuals, were cultured at the bottom of transwells (Costar 3396) together with CD4+ T cells, sorted from young individuals, placed at the top of transwells. The ratio B:CD4+ T cells was reflecting the ratio of these cell types in ex vivo isolated PBMC. After 24 hrs, CD4+ T cells were harvested, mRNA extracted and qPCR performed. Supernatants were also collected after 48 hrs for cytokine and lactate secretion measured by CBA and ELISA, respectively.

In some co-cultures, B cells were left untreated or were pre-treated overnight with the small molecule FX11 [3-dihydroxy-6-methyl-7-(phenylmethyl)-4-propylnaphthalene-1-carboxylic acid] that inhibits the enzyme LDHA (EMD Millipore 427218), or with a polyclonal antibody blocking the lactate transporter SLC5A12 (ThermoFisher Invitrogen PA5-110389), in order to block lactate secretion, before CD4+T cells were added to the top of the transwell. Concentration of FX11 was 10 mM/10^6^ B cells, whereas the anti-SLC5A12 antibody was diluted 1:500.

### Flow cytometry

PBMC (2×10^6^/mL) were membrane stained for 20 min at room temperature with Live/Dead kit and with the following antibodies: anti-CD45 (Biolegend 368540), anti-CD19 (BD 555415), anti-CD27 (BD 555441) and anti-IgD (BD 555778) to measure naive (IgD+CD27-), IgM memory (IgD+CD27+), switched memory (IgD-CD27+), and DN (IgD-CD27-) B cells.

For the evaluation of glucose uptake, cytosolic ROS, mitochondrial ROS and mitochondrial mass, PBMC were first stained with the metabolic reagents and then with Live/Dead kit and the antibodies above (anti-CD45/CD19/CD27/IgD). For glucose uptake we used with the fluorescent glucose analog (2-(N-(7-Nitrobenz-2-oxa-1,3-diazol-4-yl)Amino)-2-Deoxyglucose) (2-NBDG, Thermo Fisher N13195). For cytosolic ROS we used CellROX^®^ Deep Red Reagent (Thermo Fisher C10422), whereas for mitochondrial ROS we used with MitoSOX^TM^ Red (mitochondrial superoxide indicators, Thermo Fisher M36009). For mitochondrial mass we used MitoTracker Green (Thermo Fisher M7514). Working concentrations were recommended by the manufacturers. Staining was at room temperature for 30 minutes. Cells were then washed twice with FACS buffer, and then membrane stained for 20 minutes at room temperature. For β-Galactosidase staining, we used the Cellular Senescence detection kit SPiDER β-Gal (Dojindo Molecular Technologies SG04), but after membrane staining. In every experiment we acquired up to 10^5^ events in the B cell gate on a LSR-Fortessa (BD). Results were analyzed using FlowJo 10.5.3 software. Single color controls were included in every experiment for compensation. Isotype controls were also used in every experiment to set up the gates.

### Sorting of the B cell subsets

B cell subsets were sorted in a Sony SH800 cell sorter using anti-CD45, anti-CD19, anti-CD27 and anti-IgD antibodies. Cell preparations were typically >98% pure.

### Reverse Transcriptase (RT) and quantitative (q)PCR

RT reactions were performed in a Mastercycler Eppendorf Thermocycler to obtain cDNA. Briefly, 10 µl of mRNA from freshly-isolated cells, or 2 µl of total RNA (from Trizol, Ambion) at the concentration of 0.5 µg/µL, were used as template for cDNA synthesis in the RT reaction. For miR quantification, RNA was reverse transcribed in the presence of specific primers (provided together with the qPCR primers).

Five µL of cDNA were used for qPCR. Reactions were conducted in MicroAmp 96-well plates and run in the ABI 7500 machine. Calculations were made with ABI software. For calculations, we determined the cycle number at which transcripts reached a significant threshold (Ct) for target genes and GAPDH (control). The difference in Ct values between GAPDH and the target gene was calculated as ΔCt. Then the relative amount of the target gene was expressed as 2^-ΔCt^ and indicated as qPCR values. All reagents were from Thermo Fisher. Taqman primers were: GAPDH, Hs99999905_m1; Glut1/SLC2A1, Hs00892681; LDHA, Hs01378790_g1; PDHX, Hs00185790_m1; PPARGC1α, Hs00173304_m1; SLC5A12, Hs01054645; TNF, Hs01113624_g1; IL-6, Hs00985639_m1; IL-8, Hs00174103_m1; TLR2, Hs02621280_s1; TLR4, Hs00152939_m1; p16^INK4^ (CDKN2A), Hs00923894_m1; p21^CIP1/WAF1^, Hs00355782_m1; U6, 001973; miR-155, 002623; miR-16, 000391.

### Preparation of cytoplasmic and nuclear protein extracts and Western blot

Proteins were extracted from CD4+ T cells from young individuals after 48 hrs co-culture without or with B cells from elderly individuals. Cytoplasmic and nuclear protein extracts were prepared from the same numbers of cells. Briefly, cells were centrifuged in a 5415C Eppendorf microfuge (2,000 rpm, 5 min.). The pellet was resuspended in 20 μL of solution A containing Hepes 10 mM, pH 7.9, KCl 10 mM, EDTA 1.0 mM, DTT 1 mM, MgCl_2_ 1.5 mM, PMSF 1 mM, 1 tablet of protease inhibitor cocktail (Boeringer Manheim, Germany) (per 20 mL, 1mM Na_3_VO_4_ and Nonidet P-40 (0.1%), briefly vortexed and centrifuged (8,000 rpm, 5 min, 4°C). The supernatant containing the cytoplasmic extract (CE) was removed and stored at -80°C. To obtain nuclear extracts (NE), the pellet containing the nuclei was resuspended in solution B containing Hepes 20 mM, pH 7.9, EDTA 0.1 mM, DTT 1 mM, MgCl_2_ 1.5 mM, PMSF 2 mM, 1 tablet of protease inhibitor cocktail (per 20 mL), and glycerol 10%. The lysate was incubated on ice for 20 min, protein sonicated for a few seconds and centrifuged (14,000 rpm, 15 min, 4°C). Aliquots of NE were stored at -80°C. Protein contents were determined by Bradford assay ^18^. The amount of protein extracted from the same number of cells is highly reproducible (90%) from one experiment to another in both young and old mice.

CE and NE were denatured and then electro-transferred onto nitrocellulose filters. Filters were incubated with the following primary antibodies in PBS-Tween 20 containing 5% milk: mouse monoclonal UBC9 (1:1000 diluted, BD Bioscience 610748), mouse monoclonal NF-kB p65 (1:200 diluted, Santa Cruz Biotechnology sc-8008). After overnight incubation with the primary antibodies, immunoblots were incubated with the secondary antibody goat anti-mouse IgG (1:50,000 diluted, Jackson ImmunoResearch Labs 115-035-003) for 3 hrs at room temperature. Membranes were developed by enzyme chemiluminescence and exposed to CL-XPosure Film (Pierce). Films were scanned and analyzed using the AlphaImager Enhanced Resolution Gel Documentation & Analysis System (Alpha Innotech, San Leandro CA) and images were quantitated using the AlphaEaseFC 32-bit software.

### Mitostress test

The metabolic profile of unstimulated B cells from young and elderly individuals was evaluated by the mitostress test and Seahorse technology that allows real-time evaluation of changes in oxygen consumption rates (OCR) and extracellular acidification rates (ECAR), measures of oxidative phosphorylation and of anaerobic glycolysis, respectively. The mitostress test was conducted in a Seahorse XFp extracellular flux analyzer (Agilent). Briefly, B cells, at the concentration of 2.5×10^5^/well, were initially incubated in Seahorse XF DMEM Medium (Agilent 103575) supplemented with 2 mM glutamine (Agilent 103579), 10 mM glucose (Agilent 103577) and 1 mM pyruvate (Agilent 103578), (200 μL of each reagent in 20 mL of medium). Maximal respiratory capacity was measured by treating with Oligomycin (1 μM) to block ATP production, followed by the uncoupling agent FCCP (fluoro-carbonyl cyanide phenylhydrazone, 5 μM), to dissipate proton gradients and allow electron transport and oxygen consumption to operate at maximal rate. This elevated OCR is suppressed by Rotenone/Antimycin (1 μM), showing that respiration is mitochondrial.

### Glycolytic test

First, B cells were incubated in Seahorse XF DMEM Medium without glucose. The first injection is a saturating concentration of glucose (10 mM).The second injection is oligomycin, at the concentration of 3.5 μM. Oligomycin inhibits mitochondrial ATP production, and shifts the energy production to glycolysis, with the subsequent increase in ECAR revealing the cellular maximum glycolytic capacity. The final injection is 2-deoxy-glucose (2-DG, 20 mM), a glucose analog, that inhibits glycolysis through competitive binding to glucose hexokinase, the first enzyme in the glycolytic pathway. The resulting decrease in ECAR confirms that the ECAR produced in the experiment is due to glycolysis.

### Statistical analyses

To examine differences between 3 groups, two-way ANOVA was used. Group-wise differences were analyzed afterwards with Bonferroni’s multiple comparisons test, with p<.05 set as criterion for significance. To examine differences between 2 groups, paired Student’s t test (two-tailed) was used. To examine relationships between variables, bivariate Pearson’s correlation analyses were performed. GraphPad Prism version 9.3.1 software was used to construct all graphs.

## Results and Discussion

### Aging induces hyper-metabolic B cells

We evaluated the metabolic status of B cells isolated from the peripheral blood of young and elderly healthy individuals. Briefly, B cells were enriched from PBMC using CD19 magnetic beads. B cells were left unstimulated because we have previously shown that the status of unstimulated B cells is associated with their capacity to respond after in vivo or in vitro stimulation. We used the mitostress test and Seahorse technology that allow real-time evaluations of oxygen consumption rates (OCR), measure of oxidative phosphorylation (OXPHOS) and mitochondrial fitness, and extracellular acidification rates (ECAR), measure of anaerobic glycolysis and lactate secretion. In the mitostress test, the sequential addition of pharmacological drugs alters the bioenergetic profile of the mitochondria. Results in Fig. 1 show higher OCR (a), higher maximal respiration (b) and higher spare respiratory capacity (c) in B cells from elderly versus young individuals. The differences in ECAR in B cells from elderly as compared to those from younger individuals are even higher than those in OCR (d), with higher maximal ECAR (e) and lower ratio OCR:ECAR under maximal respiratory conditions (f). Collectively, there findings indicate that B cells from elderly individuals are able to switch more readily from OXPHOS to anaerobic glycolysis after ATP synthase is blocked by the addition of oligomycin in the mitostress test. No differences were observed in the other measures performed during the mitostress test (non mitochondrial respiration, coupling efficiency, proton leak, ATP production) (Supplementary Fig. 1a, b, c, d).

**Fig. 1.**
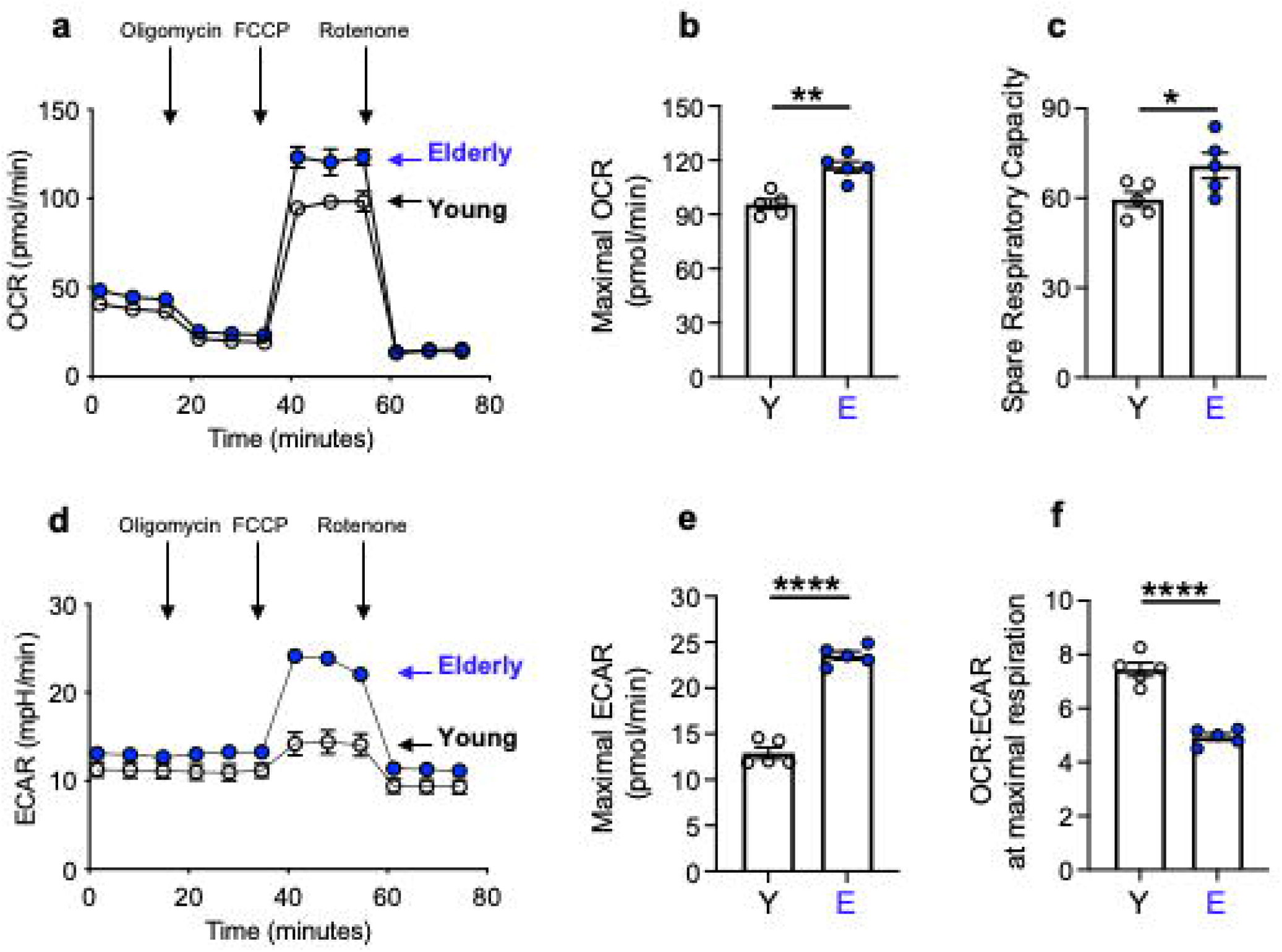
Aging induces hyper-metabolic B cells. B cells, isolated from the peripheral blood of young and elderly individuals using magnetic beads, were left unstimulated. B cells were seeded into the wells of an extracellular flux analyzer at the concentration of 2×10^5^/well in triplicate and run in a mitostress test. **a.** OCR results representative of 5 independent experiments. **b.** Maximal OCR. **c.** Spare respiratory capacity (calculated subtracting basal OCR from maximal OCR values). **d.** ECAR results representative of 5 independent experiments. **e.** Maxinal ECAR. **f.** OCR:ECAR at maximal respiration. Mean comparisons between groups were performed by unpaired Student’s t test (two-tailed). *p<0.05, **p<0.01, ****p<0.0001

### Aging induces glycolytic B cells

We wanted to confirm the above results by measuring by qPCR the expression of transcripts for enzymes involved in metabolic pathways associated with OCR and ECAR. In particular, we measured the expression of a member of the pyruvate deahydrogenase (PDH) complex, PDHX, that converts pyruvate into Acetyl CoA and therefore represents a link between glycolysis, and Kreb’s cycle and ROS production; and the expression of lactate dehydrogenase A (LDHA), that converts pyruvate into lactate. First, we measured by qPCR the expression of the transcript for glucose transporter 1 (Glut1). Fig. 2a shows higher expression of Glut1 RNA in unstimulated B cells from elderly individuals, as compared to those from younger controls. Glut1 is the major glucose transporter of human circulating B cells that also express transcripts for Glut4 albeit at lower levels, whereas transcripts for Glut2 and Glut3 are expressed at extremely low levels and no age differences were detected (Supplementary Fig. 2a, b, c, d, e). We also measured glucose uptake by flow cytometry and the glucose fluorescent analog [2-(N-(7-Nitrobenz-2-oxa-1,3-diazol-4-yl)Amino)-2-Deoxyglucose], 2-NBDG. Fig. 2b, c shows higher glucose uptake in unstimulated B cells from elderly versus young individuals, confirming the qPCR results on Glut1 expression.

**Fig. 2.**
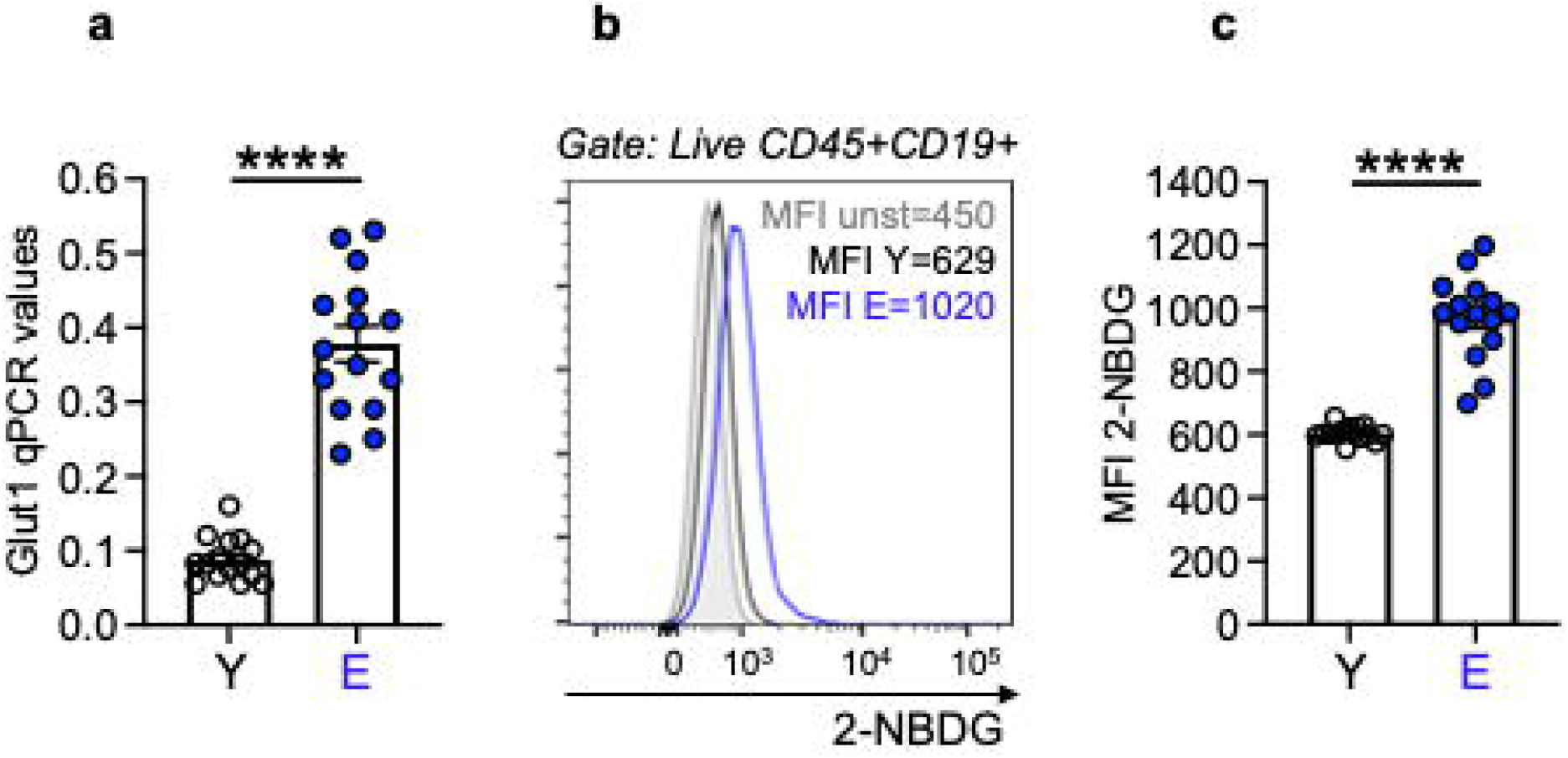
Aging induces higher glucose uptake in unstimulated B cells. B cells, isolated as in Fig. 1, were left unstimulated. **a.** mRNA expression of the glucose transporter Glut1 measured by qPCR. Results show qPCR values (2^-ΔCt^). **b.** Representative flow cytometry histograms of glucose uptake measured by flow cytometry using the glucose fluorescent analog 2-NBDG. Results show MFI (mean fluorescence intensity). **c.** 2-NBDG MFI results from all individuals. Mean comparisons between groups were performed by unpaired Student’s t test (two-tailed). ****p<0.0001

We then evaluated PDHX RNA expression and found it higher in unstimulated B cells from elderly versus young individuals (Fig. 3a), as well as the expression of cytosolic ROS (reactive oxygen species), that reflects almost exclusively mitochondrial ROS, as evaluated by flow cytometry and CellROX staining (Fig. 3b, c). Mitochondrial ROS was also measured in some individuals by flow cytometry and MitoSOX staining, with results showing similar age differences as those above (Fig. 3d). Moreover, B cells from elderly individuals show comparable total mitochondrial mass as compared to those from young controls, as evaluated by flow cytometry and the mitochondrial probe MitoTracker Green (Fig. 3e, f), but increased mRNA expression of the peroxisome proliferator-activated receptor-γ coactivator 1α (PPARGC1α) (Fig. 3g), a co-transcriptional regulator involved in mitochondrial biogenesis ^19^.

**Fig. 3.**
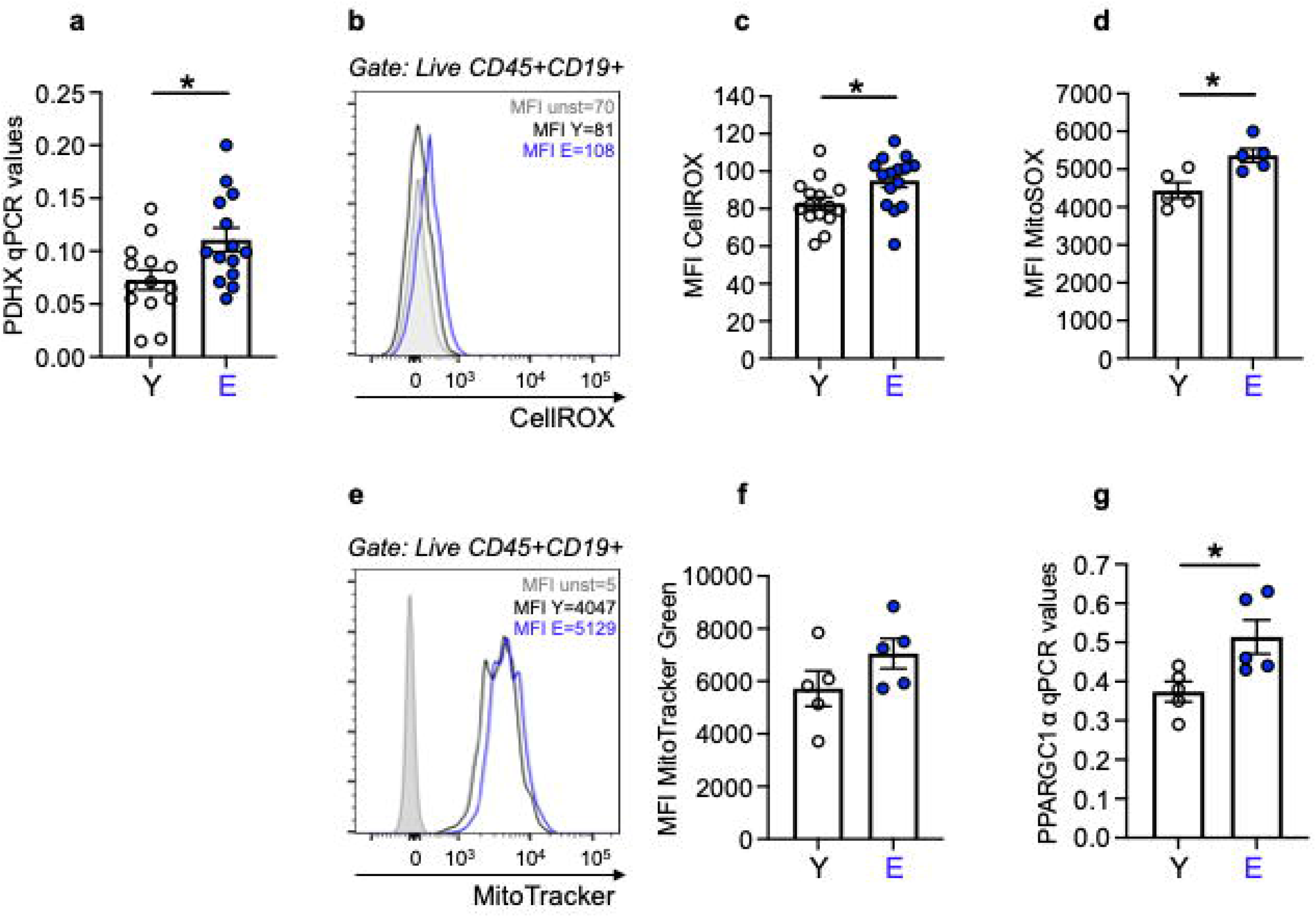
Aging increases mitochondrial function in unstimulated B cells. B cells, isolated as in Fig. 1, were left unstimulated. **a.** mRNA expression of PDHX. Results show qPCR values (2^-ΔCt^). **b.** Representative flow cytometry histograms of cytoplasmic ROS staining using the fluorescent compound CellROX. **c.** Cytoplasmic ROS MFI results from all individuals. **d.** Mitochondrial ROS MFI results from all individuals after staining with the fluorescent compound MitoSOX. **e.** Representative flow cytometry histograms of mitochondrial mass staining using the MitoTracker Green probe. **f.** MitoTracker Green MFI results from all individuals **g.** mRNA expression of PPARGC1α. Results show qPCR values (2^-ΔCt^). Mean comparisons between groups were performed by unpaired Student’s t test (two-tailed). *p<0.05

Although PDHX RNA expression and ROS production were significantly higher in unstimulated B cells from elderly as compared to those from younger individuals, the biggest difference was observed when we evaluated RNA expression of LDHA (Fig. 4a) and lactate secretion (Fig. 4b). LDHA transcripts were evaluated in unstimulated B cells whereas lactate secretion was quantified by ELISA after B cell stimulation for 48 hrs with the mitogen CpG. Autoimmune antibody secretion was also evaluated in 7-day CpG-stimulated B cells (Fig. 4c), and double strand (ds)DNA-specific IgG autoimmune antibodies were positively and significantly associated with lactate (Fig. 4d). These results are to our knowledge the first to show that lactate induces autoimmune pathogenic B cells and is a stimulus for autoimmune antibody secretion. Other autoimmune specificities were measured (Supplementary Fig. 3).

**Fig. 4.**
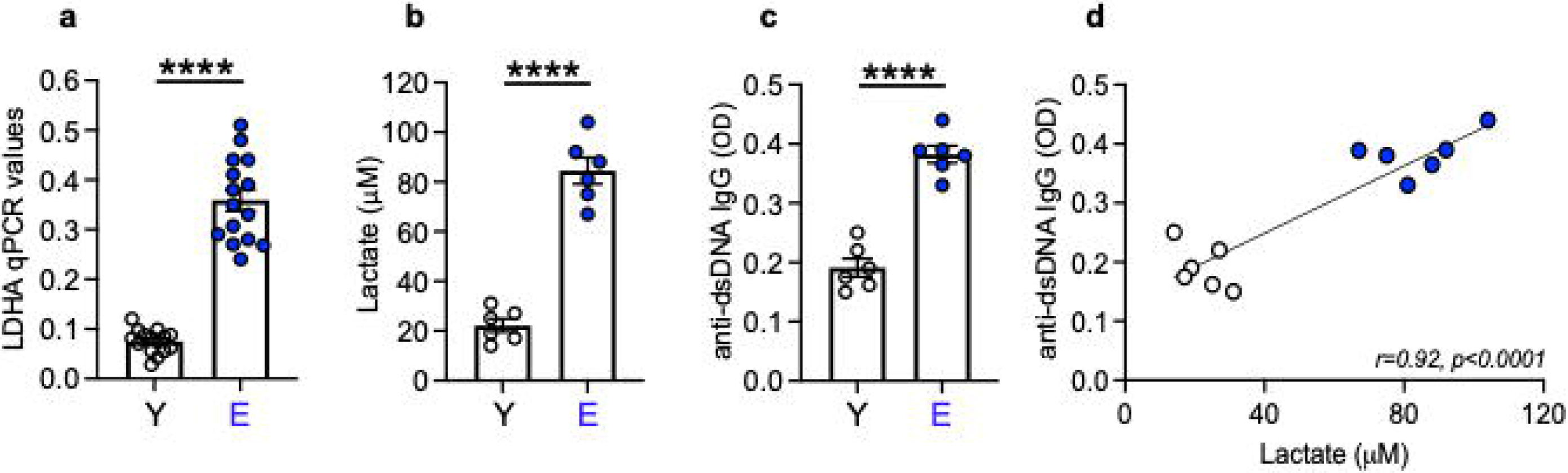
Aging increases anaerobic glycolysis in unstimulated B cells. B cells, isolated as in Fig. 1, were left unstimulated. **a.** mRNA expression of LDHA. Results show qPCR values (2^-ΔCt^). **b.** Lactate secretion in culture supernatants of CpG-stimulated B cells (48 hrs) measured by ELISA. **c.** Anti-dsDNA autoimmune IgG antibodies in culture supernatants of CpG-stimulated B cells (7 days) measured by ELISA. **d.** Correlation of lactate and autoimmune IgG. Pearson’s r and p values are shown at the bottom of the figure. Mean comparisons between groups were performed by unpaired Student’s t test (two-tailed). ****p<0.0001.

### B cells from elderly individuals induce pro-inflammatory CD4+ T cells through metabolic pathways

We then evaluated if hyper-metabolic B cells from elderly individuals were also capable to polarize CD4+ T cells from young individuals making them hyper-metabolic, pathogenic and inflammatory. To do so, we set up co-cultures in which CD4+ T cells from young individuals were positioned in the top part of a transwell, whereas B cells from young or elderly individuals were positioned at the bottom of the transwell (Supplementary Fig. 4a). After 24 hrs, CD4+ T cells were harvested and evaluated by qPCR for the expression of transcripts for Glut1, LDHA and PDHX, whereas after 48 hrs, supernatants were tested for the presence of T cell-derived pro-inflammatory cytokines IL-17A and IFN-γ. Fig. 5a shows that CD4+ T cells from young individuals were able to up-regulate the expression of transcripts for Glut1 and LDHA, and to a lesser extent for PDHX, only in the presence of B cells from elderly individuals, whereas no effect was observed if B cells were from young individuals. Fig. 5b shows that supernatants from co-cultures of young CD4+ T cells with B cells from elderly but not young individuals were enriched in the T cell-derived pro-inflammatory cytokines IL-17A and IFN-γ at 48 hrs. In these supernatants other pro-inflammatory cytokines (IL-6, IL-10, TNF-α) were also enriched but to a lesser extent (Supplementary Fig. 4b, c, d), whereas no changes were observed for IL-2 and IL-4 (Supplementary Fig. 4e, f). The same supernatants were also tested for the presence of lactate (Fig. 5c). We found lactate only in co-cultures set up in the presence of B cells from elderly individuals. In these conditions, lactate comes exclusively from B cells, as CD4+ T cells need to be heavily stimulated with the mitogens PMA and Ionomycin to secrete lactate (data not shown). Moreover, the amount of lactate in these co-cultures is much less that that measured in CpG-stimulated B cells (see Fig. 4b).

**Fig. 5.**
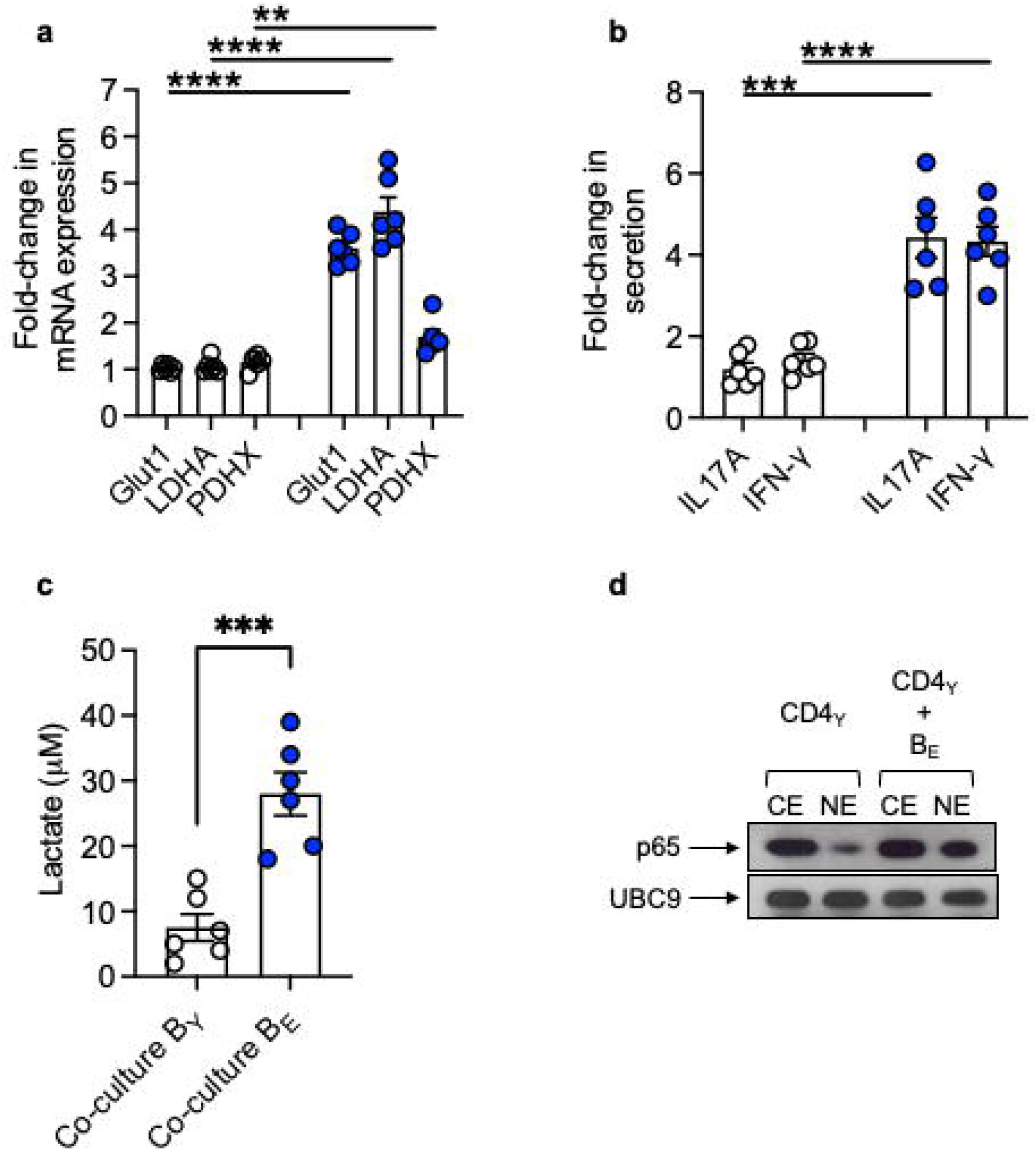
B cells from elderly individuals induce pro-inflammatory CD4+ T cells through metabolic and inflammatory pathways. B cells from young (white symbols) and elderly (blue symbols) individuals were cultured at the bottom of transwells together with CD4+ T cells, sorted from young individuals, placed at the top of transwells. After 24 hrs, CD4+ T cells were harvested, mRNA extracted and qPCR performed. **a.** Fold-change in RNA expression of metabolic markers (Glut1/LDHA/PDHX). **b.** Fold-change in IL-17A and IFN-γ secretion measured by CBA. **c.** Lactate measured after 48 hrs by ELISA. **d.** Cytoplasmic (CE) and nuclear (NE) protein extracts (5 μg/lane) from CD4+ T cells from young individuals, alone or in co-culture with B cells from elderly individuals, were blotted with the indicated antibodies. Results are representative of 2 independent experiments. Mean comparisons between groups were performed by unpaired Student’s t test (two-tailed). **p<0.01, ***p<0.001, ****p<0.0001.

CD4+ T cells from young individuals co-cultured with B cells from elderly individuals show NF-kB activation, measured by the translocation from the cytoplasm to the nucleus of RelA/p65, as compared to those that are not co-culted with B cells (Fig. 5d), suggesting that the lactate induction of pro-inflammatory cytokine secretion occurs through NF-kB, as also previously shown in our mouse experiments ^20^.

Experiment in progress in the laboratory have supported the polarizing effect on B cells from elderly individuals not only on CD4+ T cells but also on CD8+ T cells and monocytes from young individuals (data not shown). Similar to the results above, the mechanisms indicate lactate and NF-kB as crucial players.

We next tried to block lactate using specific inhibitors and evaluate if blocking lactate was also reducing IL-17A and IFN-γ secretion as shown in Supplementary Fig. 4g, h. We used two different lactate inhibitors: FX11, [3-dihydroxy-6-methyl-7-(phenylmethyl)-4-propylnaphthalene-1-carboxylic acid], a small molecule that blocks the enzyme LDHA, and anti-SLC5A12, a polyclonal antibody that inhibits the lactate transporter SLC5A12. This transporter acts as an electroneutral and low affinity sodium-dependent solute transporter that catalyzes the transport across the membrane of many monocarboxylates including lactate. Fig. 6a shows that pre-inclubation of B cells from elderly individuals with FX11 before the co-culture significantly inhibits the secretion of both cytokines. However, when we used the anti-SLC5A12 antibody the inhibition was even more effective and the two cytokines were barely detectable in culture supernatants (Fig. 6b).

**Fig. 6.**
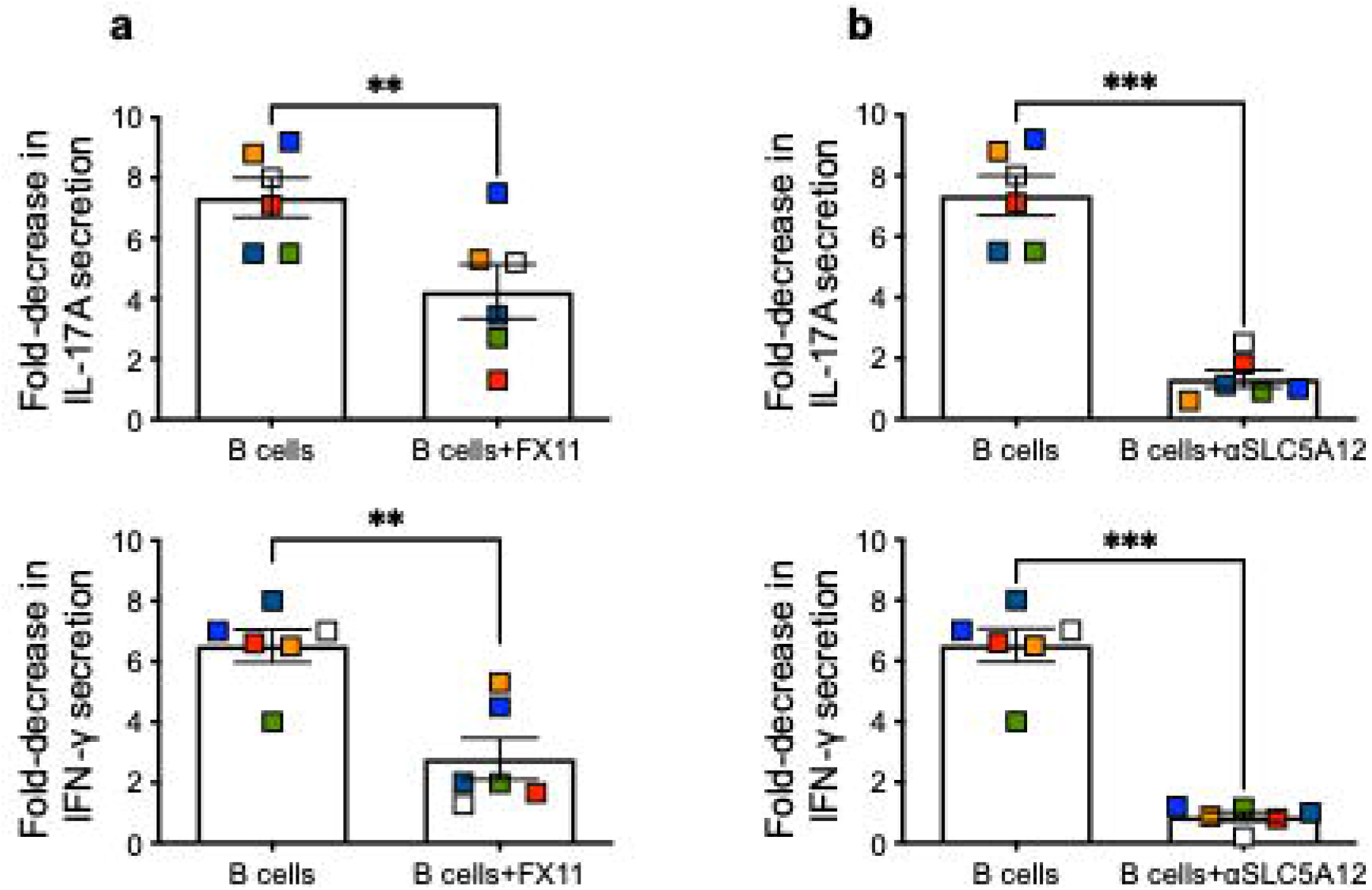
Lactate inhibition decreases the polarizing effect of B cells from elderly individuals. B cells from elderly individuals were cultured at the bottom of transwells together with CD4+ T cells from young individuals, placed at the top of transwells. B cells were left untreated or pre-treated overnight with FX11 (10 mM/10^6^ B cells) (**a**) or with anti-SLC5A12 (1:500 dilution) (**b**), inhibitors of LDHA or of the lactate transporter SLC5A12, respectively, before CD4+ T cells were added at the top of the transwell. Supernatants were collected after 48 hrs to measure IL-17A and IFN-γ secretion by CBA. Results show fold-decrease in secretion of the cytokines. Mean comparisons between groups were performed by paired Student’s t test (two-tailed). **p<0.01, ***p<0.001.

### Lactate induces immunosenescent B cells that are hyper-glycolytic and hyper-inflammatory

Next, we wanted to examine if lactate was also able to induce pro-inflammatory B cells, similar to what we have described above for CD4+ T cells. Briefly, B cells from young individuals were stimulated overnight with GpG in the presence or absence of lactate. Then, cultures were harvested and cells loaded on a glycolytic test in Seahorse performed with injections of glucose, olygomycin and 2-DG. Fig. 7a shows that lactate induces a hyper-glycolytic phenotype in B cells from young individuals which is confirmed by expression of Glut1 and LDHA transcripts and, to a lesser extent, of PDHX (Fig. 7b). B cells stimulated overnight in the presence of lactate look indistinguishable from B cells from elderly individuals, clearly indicating that lactate induces immunosenescece in B cells from young individuals. Lactate increases expression of transcripts for multiple inflammatory markers, many of which are members of the SASP, Fig. 7c. These include pro-inflammatory cytokines and chemokines (TNF, IL-6, IL-8), pro-inflammtory microRNAs (miRs) involved in class switch and antibody secretion (miR-155, miR-16), markers of immune activation (TLR2, TLR4) and cell cycle inhibitors (p16^INK4^, p21^CIP1/WAF1^). In line with the expression of SASP transcripts, B cells from young individuals overnight stimulated with lactate were positively stained with the marker of immunosenescence β-Galactosidase, similar to B cells from elderly individuals (Fig. 7d). After 48 hrs of CpG stimulation in the presence of lactate, B cells from young individuals, similar to those from elderly individuals, secrete IL-6 and TNF-α in culture supernatants (Fig. 7e, f), and after 7 days also secrete autoimmune pathogenic antibodies (Fig. 7g).

**Fig. 7.**
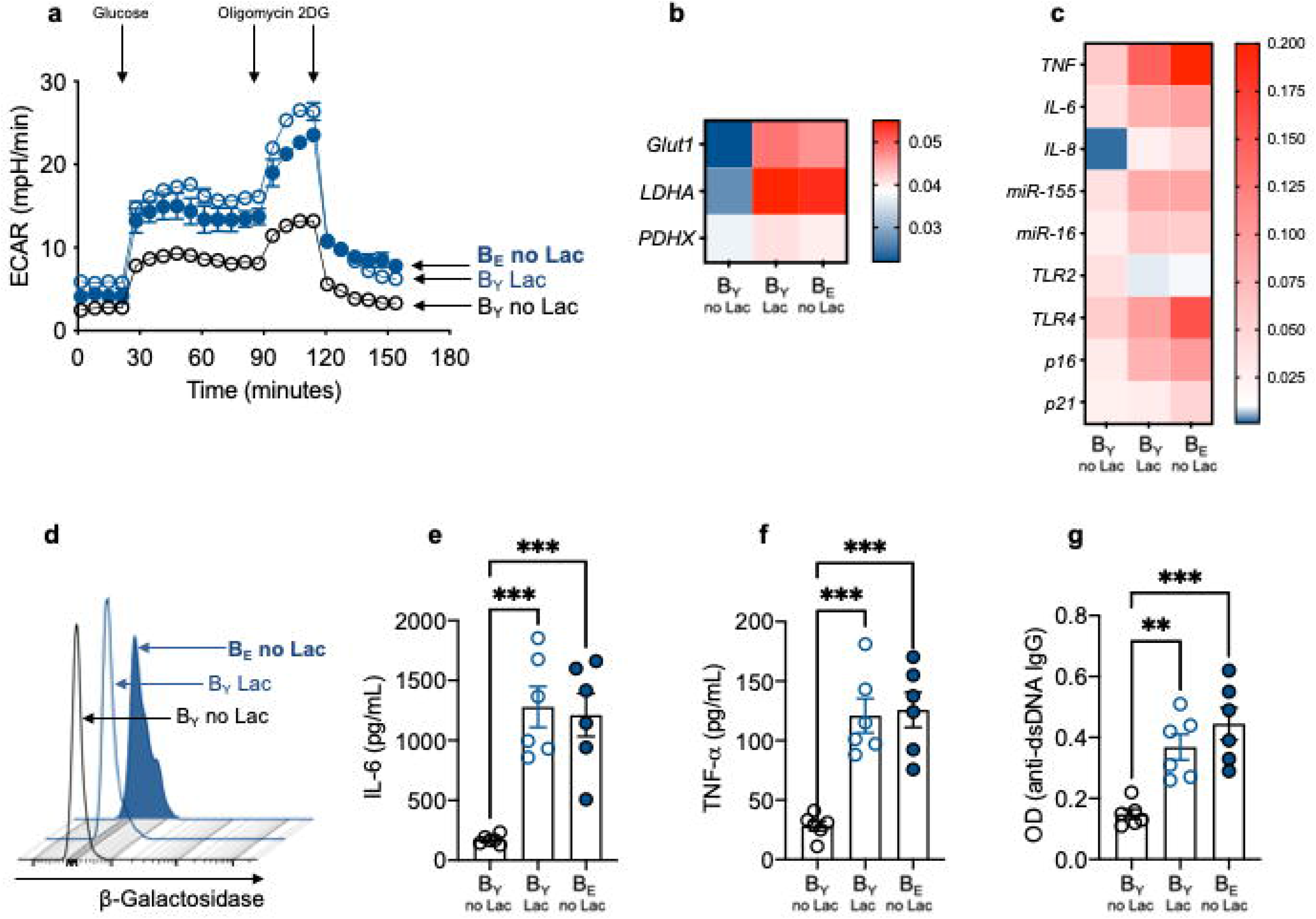
Lactate induces immunosenescent B cells that are hyper-glycolytic and hyper-inflammatory. B cells from young and elderly individuals, isolated as in Fig. 1, were stimulated overnight with CpG (5 µg/10^6^ cells). B cells from young individuals were also stimulated with CpG+lactate (10 mM/10^6^ B cells). **a**. Cells were seeded into the wells of an extracellular flux analyzer at the concentration of 2×10^5^/well and run in a glycolytic test. **b**. Expression of transcripts for metabolic markers. **c**. Expression of transcripts for pro-inflammatory markers. Heapmaps in **b** and **c** show qPCR values (2^-ΔCt^) of multiple pro-inflammatory markers, normalized to GAPDH. **d**. Staining with β-Galactosidase. Results show MFI from one representative experiment (values from all individuals are: 130±5, young; 243±29, young+lactate; 296±35, elderly no lactate). **e**. IL-6 in culture supernatants after 48 hrs measured by CBA. **f.** TNF-α in culture supernatants after 48 hrs measured by CBA. **g**. Autoimmune IgG antibodies after 7 days measured by ELISA. Mean comparisons were performed by two-way ANOVA. ***p<0.001.

The effects of lactate on B cells from elderly individuals occur because B cells express the lactate tranporter SLC5A12, as shown in Supplementary Fig 5a. The expression is low but detectable on CpG-stimulated B cells from young individuals and is up-regulated by the overnight stimulation in the presence of lactate. In this condition, SLC5A12 expression is comparable to what is observed in CpG-stimulated B cells from elderly individuals. SLC5A12 expression can be effectively inhibited after a short incubation of B cells with the neutralizing antibody specific for the transporter, and higher is the expression of SLC5A12 higher is the level of inhibition (Supplementary Fig 5b). In the presence of the anti-SLC5A12 neutralizing antibody, secretion of IL-6 and TNF-α as well of autoimmune pathogenic antibodies is inhibited in B cells from young individuals incubated in the presence of lactate (Fig. 8a, b, c), and in B cells from elderly individuals (Fig. d, e, f).

**Fig. 8.**
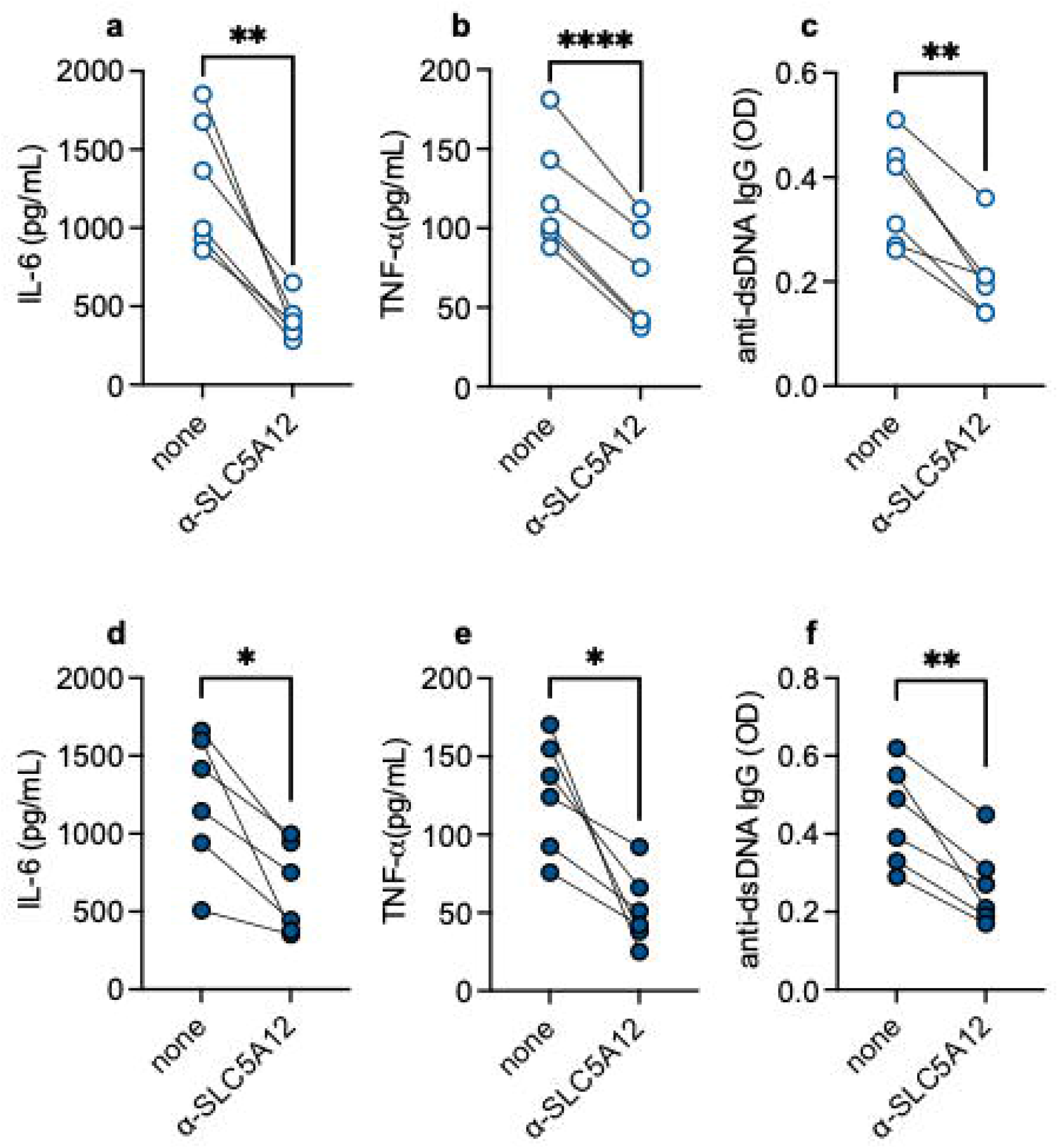
The effects of lactate on B cells from young individuals are decreased by inhibition of the lactate transporter. B cells from young and elderly individuals, isolated as in Fig. 1, were stimulated overnight with CpG (5 µg/10^6^ cells). B cells from young individuals were stimulated with CpG in the presence of lactate. Secretion of IL-6 (**a**), TNF-α (**b**) and autoimmune IgG (**c**) in cultures of B cells from young individuals+lactate or from elderly (**d-f**) individuals. Mean comparisons between groups were performed by paired Student’s t test (two-tailed). *p<0.05, **p<0.001, ****p<0.0001.

### The hyper-metabolis status of B cells from elderly individuals reflects the age-associated expansion of pro-inflammatory B cell subsets

We have previously shown that the composition of the circulating B cell pool changes with age with a significant expansion of the subset of Double Negative (DN) B cells which is the most pro-inflammatory B cell subset ^21–23^. Therefore, we wanted to investigate the metabolic status of DN B cells as compared to that of the other circulating B cell subsets. The composition of the circulating B cell pool of the recruited young and elderly individuals is shown in Supplementary Fig. 6. Fig. 9 shows that memory B cells are more metabolic than naïve B cells, in both young and elderly individuals, and that among memory B cells DN B cells express the highest levels of transcripts for Glut1 (a), LDHA (b) and also PDHX (c). We then compared the expression of transcripts not only for Glut1 and LDHA but also for many other enzymes involved in anaerobic glycolysis (Supplementary Fig. 7) in DN sorted for young and elderly individuals, and found all of them significantly higher in elderly versus young DN B cells (d). These results are the first to our knowledge to show that the pro-inflammatory phenotype of DN B cells is metabolically supported, and mainly by pathways of anaerobic glycolysis.

**Fig. 9.**
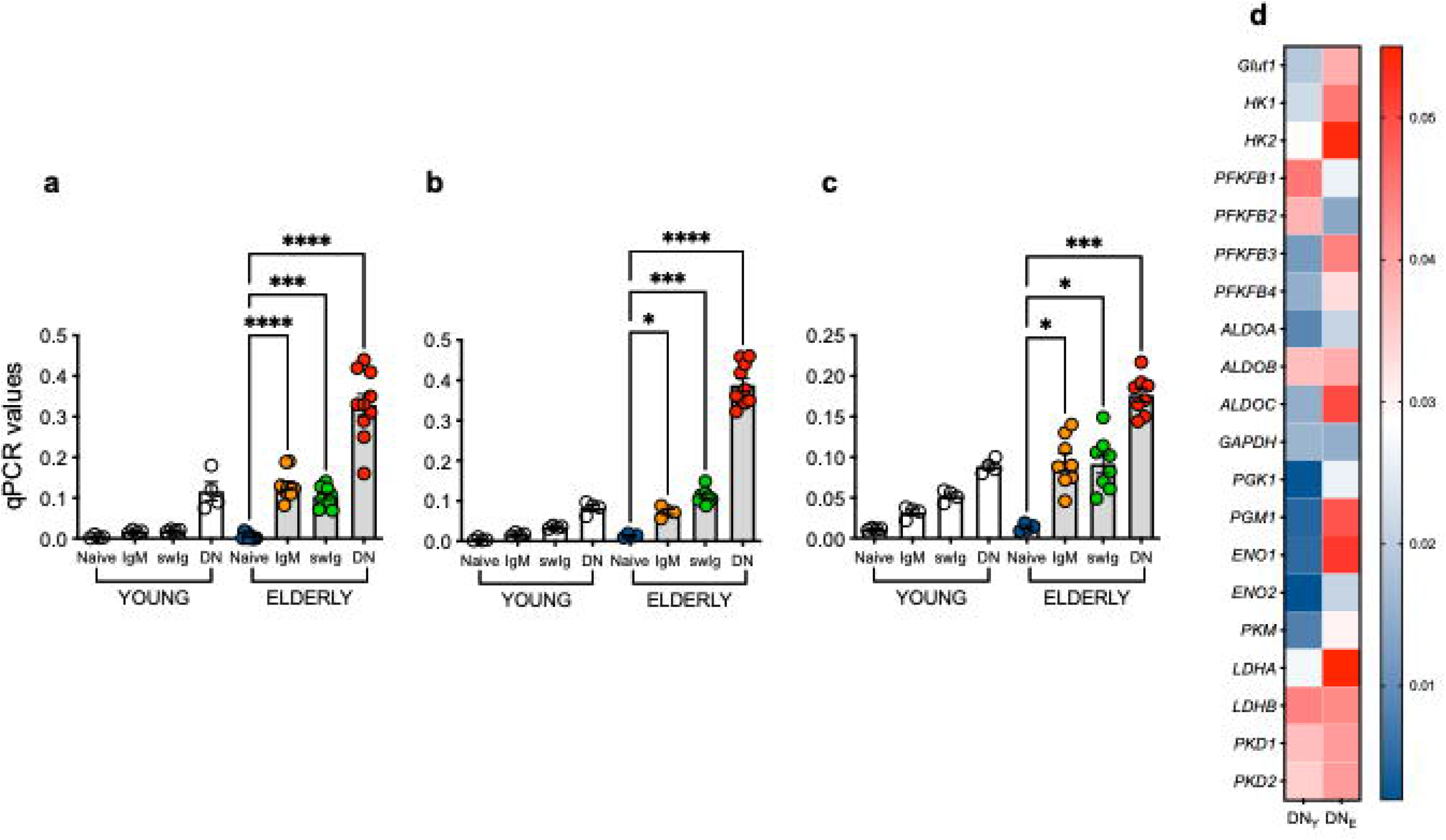
The hyper-metabolis status of B cells from elderly individuals reflects the age-associated expansion of pro-inflammatory B cell subsets. The four major circulating B cells subsets (naïve, IgM memory, swIg and DN B cells) were sorted from young and elderly individuals and left unstimulated. The mRNA was extracted at the end of sorting to evaluate the expression of transcripts for the metabolic markers Glut1 (**a**), LDHA (**b**), PDHX (**c**) by qPCR. **d.** Heapmap shows qPCR values (2^-ΔCt^) of multiple transcripts for glycolytic enzymes, normalized to GAPDH. Mean comparisons between groups were performed by paired Student’s t test (two-tailed). *p<0.05, ***p<0.001, ****p<0.0001.

## Data Availability

The data and materials that support the findings of this study are available upon request to the corresponding author.

## Acknowledgements

The authors would like to thank the volunteers who participated in this study, and the personnel of the Department of Family Medicine and Community Health at the University of Miami Miller School of Medicine, in particular Dr. Robert Schwartz, Chairman, for the recruitment of healthy participants. The authors also thank Dr. Bonnie Blomberg for sharing lab space and reagents.

## Author contributions

DF wrote the paper. DF, MR, KM, DG, AG, KL, DS performed the experiments and analyzed data. All authors have seen and approved the manuscript. DF was involved in funding acquisition.

