## Supplementary figures and images for "Immunometabolic effects of lactate on B cell function in healthy individuals of different ages"

### Supplemental Fig.1

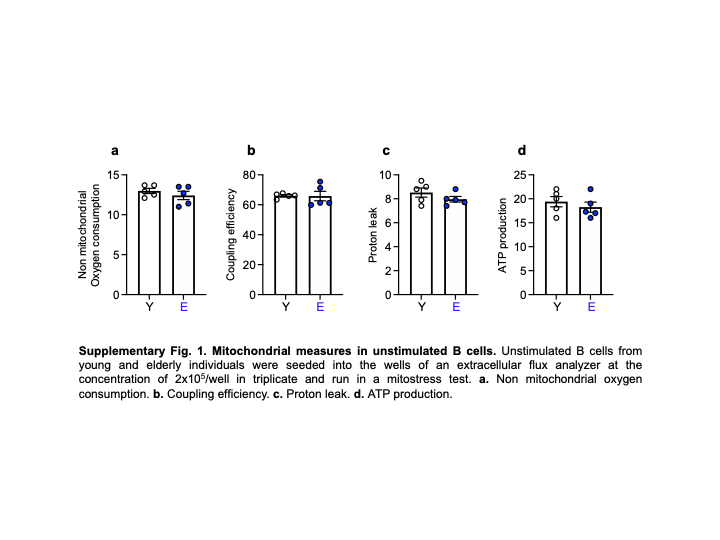

### Supplemental Fig.2

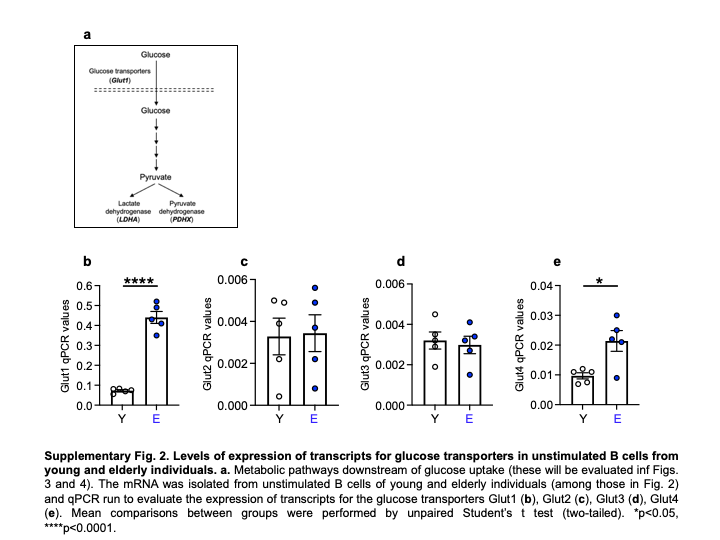

### Supplemental Fig.3

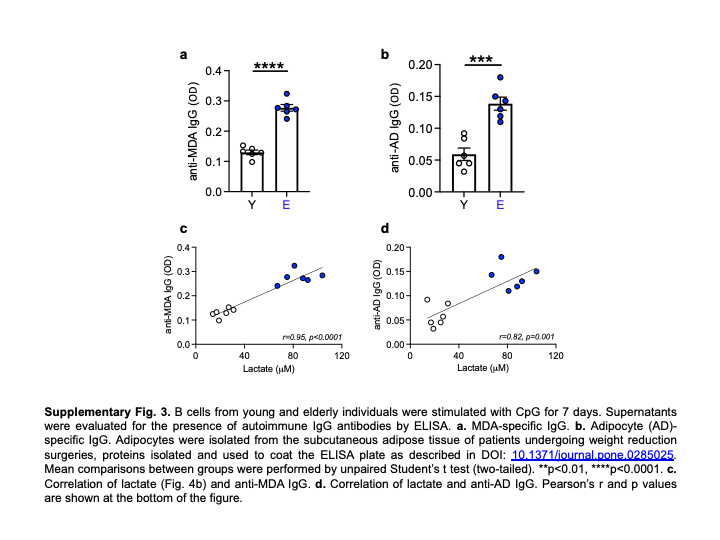

### Supplemental Fig.4

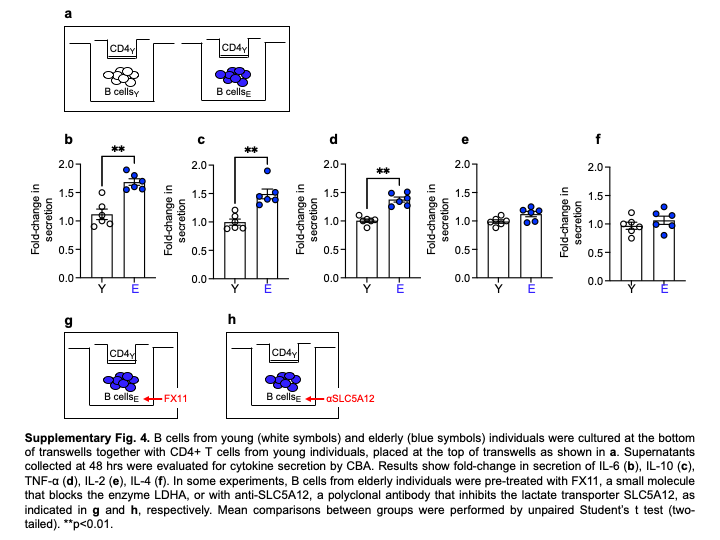

### Supplemental Fig.5

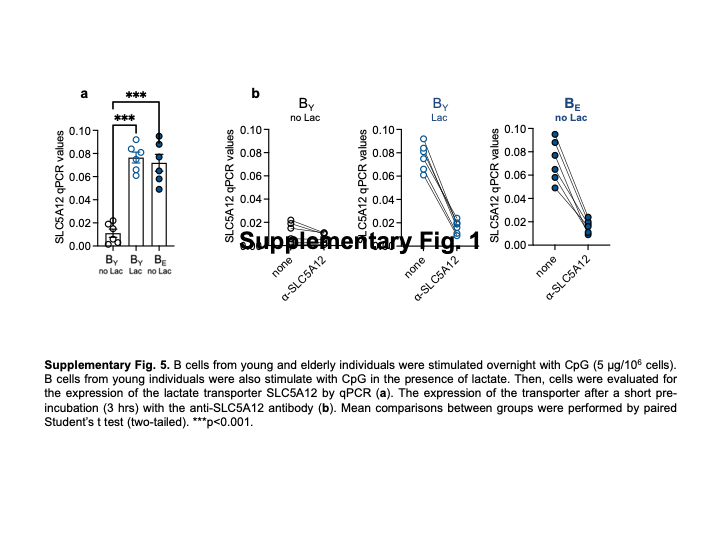

### Supplemental Fig.6

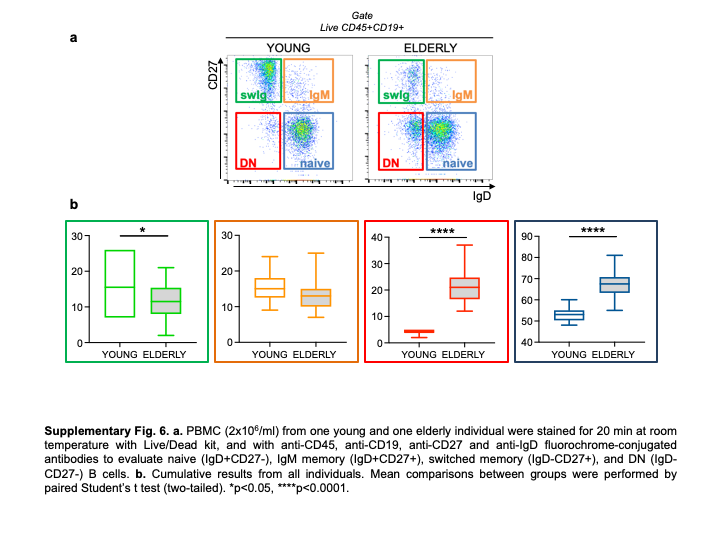

### Supplemental Fig.7

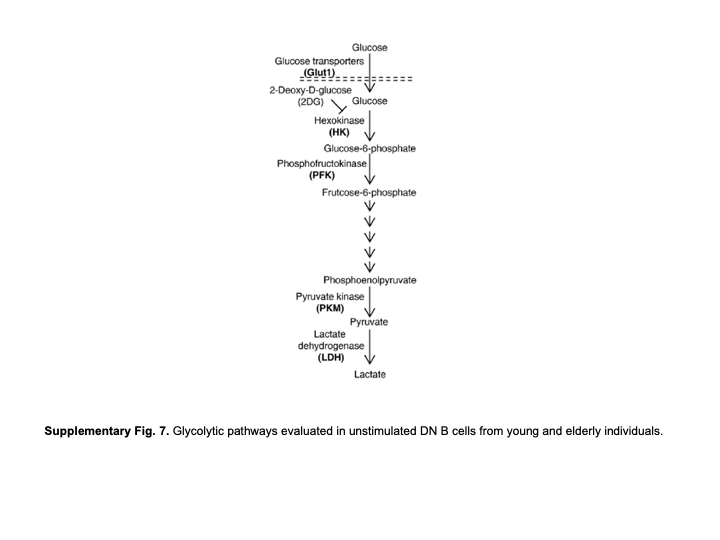
