## Supplemental Table 1 for "Immunometabolic effects of lactate on B cell function in healthy individuals of different ages"

**Supplementary Table 1.** Characteristics of the recruited participants (age, gender, serum metabolic measures)

|  | YOUNG | ELDERLY |
| --- | --- | --- |
| Age | 30-45 | ≥65 |
| Males | 11 | 10 |
| Females | 9 | 10 |
| Low-Density Lipoprotein (LDL), mg/dL | 92±1 | 104±4** |
| High-Density Lipoprotein (HDL), mg/dL | 63±2 | 58±163** |
| Glucose, mg/dL | 75±2 | 99±2**** |
| C-reactive protein (CRP), mg/L | 1.4±0.03 | 3.1±0.1**** |
| Serum Amyloid A (SAA), mg/L | 2±0.1 | 5±0.4**** |
| Lactate Dehydrogenase (LDH), U/L | 145±5 | 257±13**** |
| Non-esterified Fatty Acids (NEFA), nMol/mL | 159±9 | 217±17** |

**p<0.01

****p<0.0001 (unpaired Student’s t test)
